# Risk Factors for Cerebral Palsy in Brazilian Children: A Case-Control Study

**DOI:** 10.1101/2020.12.01.20242248

**Authors:** Marcus Valerius da Silva Peixoto, Andrezza Marques Duque, Allan Dantas dos Santos, Shirley Verônica Melo Almeida Lima, Caíque Jordan Nunes Ribeiro, Silvia Maria Voci, Susana de Carvalho, Marco Antônio Prado Nunes

## Abstract

**Background:** Cerebral palsy is the main cause of physical disability in childhood.

**Objectives:** This study analyzed prenatal and perinatal risk factors that contribute to cerebral palsy in Brazilian children.

**Methods:** A case-control study was conducted with 2- to 10-year-old children in the city of Aracaju, Sergipe, Brazil. The cases were population-based, selected from the Primary Health Care services. The controls were selected from the database of the Brazilian Live Births Information System. Controls were paired with cases by gender, year, and hospital of birth.

**Results:** A total of 570 participants (114 cases and 456 controls) were studied. Most of the participants were male, with bilateral spastic cerebral palsy. Among the prenatal factors examined, the presence of congenital anomalies was significantly different between cases and controls (OR = 54.28, [95% CI 12.55, 234.86]). The analysis of perinatal factors revealed significant differences between cases and controls in low birth weight (OR = 3.8, [95% CI 2.34, 6.16]), preterm birth (OR = 2.31, [95% CI 1.41, 3.80]), and low Apgar scores (OR = 14.73, [95% CI 5.27, 41.15]).

**Conclusions:** The main prenatal and perinatal factors associated with cerebral palsy in our population were congenital anomalies, low Apgar scores, low birth weight, and preterm birth. The perinatal period had more risk factors, demanding a deeper study of their causes and of possible preventive measures.

## INTRODUCTION

Cerebral palsy (CP) is the leading cause of childhood physical disability. It encompasses a group of permanent movement and postural deficiencies that impair activity, and are caused by non-progressive injuries that occur in the development of the fetal or infant brain. Motor impairment from cerebral palsy may also be accompanied by primary or secondary disturbances of sensation, perception, cognition, communication, and behavior, by epilepsy, and by secondary musculoskeletal problems.^1,2^

The known risk factors for cerebral palsy in full-term children in developed countries are placental abnormalities, congenital malformations, low birth weight, meconium aspiration syndrome, emergency cesarean delivery (C-section), asphyxia during delivery, neonatal seizures, respiratory distress syndrome, hypoglycemia, and neonatal infections.^3,4^

Small for gestational age (SGA) and low birth weight may be considered as the main risk factors for cerebral palsy.^2,5^ In addition to the multifactorial clinical aspects, the causes of CP can also be related to healthcare management and quality.^6^

A systematic review reported a prevalence of 2.11 per 1,000 live births.^7^ The low prevalence of CP makes it difficult to identify risk factors. However, case-control studies provide a methodological tool that can contribute to their understanding.^8^

Most of the epidemiological studies on CP were conducted in developed countries,^4,7,9,10^ despite efforts by researchers from various developing countries.^6,11–16^ An increased risk of cerebral palsy in low- and middle-income countries, due to exposure to perinatal events, has been suggested.^2,17^ The epidemiological aspects of CP need further clarification, especially in socioeconomic contexts characterized by social inequities.

The goal of this study is to analyze prenatal and perinatal risk factors contributing to cerebral palsy in Brazilian children.

## METHODS

### Study Design

A case-control study was conducted in the city of Aracaju, Sergipe, Brazil, organized in two stages: a) Identification of CP cases in a population-based survey of the whole city; b) Search of prenatal and birth information on cases and controls in the records of the Brazilian Ministry of Health Live Births Information System (SINASC).

### Study area

According to the Brazilian Institute of Geography and Statistics (IBGE), Aracaju has a surface of 181.90 km^2^, an estimated population of 623,766, and no rural areas.

The city provides primary health care coverage for 93% of the population, with 44 health units and 144 primary health care teams, each team including medical, nursing, and community health professionals. Hospital care for pregnant women is provided by two maternity hospitals.

### Data Collection

#### Cases

The cases were identified in the early stages of the research, through an active search in the 44 primary health care units of the city. Primary care teams collaborated as key informants, based on their knowledge of their respective geographical sectors. After the identification of the cases, a questionnaire was administered to the biological mothers of children with CP, at their homes. Gestational history and childbirth information were gathered from the SINASC in the second stage of the research.

##### Inclusion criteria

Children 0–2 years old, born between 2006 and 2016, living in Aracaju, with a diagnosis of cerebral palsy according to the International Classification of Diseases - ICD-10 (G.80.0-9).^18^

##### Exclusion Criteria

Children with post-natal cerebral palsy, those with absent biological mothers, and those diagnosed with progressive encephalopathies.

#### Controls

The controls were selected at the SINASC. The cases were excluded from the database for the selection of the controls, which were automatically drawn by means of a script with the R software. The pairing was performed by gender, hospital and year of birth at the rate of 1:4.

### Variables

The Brazilian Ministry of Health Live Birth Information System collects data on 106 variables. The final analysis included 14 variables, organized as follows: *Maternal age* (adolescent mother: ≤ 19 years old; adult mother: 20–34 years old; mother age 35 and above); *maternal schooling* (≤3 years of schooling, 4–7 years of schooling, 8–11 years of schooling, ≥12 years of schooling); *mother’s marital status*; *number of children alive*; *historic of number of fetal losses and miscarriages, gestational age* (preterm: ≤36 weeks; full-term: 37–42 weeks); *single or multiple pregnancy*; *type of delivery* (natural or C-section); *number of prenatal consultations*; *child’s gender*; *Apgar 1’* (normal: ≥7; low: ≤6); *Apgar 5’* (normal: ≥7; low: ≤6); *weight at birth* (normal: ≥2500 g; low: <2500 g); and *presence or absence of congenital anomalies*.

### Data Analysis

Descriptive analysis of absolute and relative frequencies of categorical variables and measurements of central tendency and variability for numerical variables were performed. Differences between proportions were analyzed using Pearson’s chi-squared test or Fisher’s exact test, while Student’s t-test was used to compare two independent samples of numerical variables. Univariate and multivariate logistic regression was applied to identify predictive factors.

### Ethical Aspects

The research was approved by the Federal University of Sergipe’s Research Ethics Committee (opinion number “1.177.455”) which abides by the ethical principles of the Brazilian National Council of Ethics in Research (Conselho Nacional de Ética em Pesquisa do Brasil - CONEP) and the Declaration of Helsinki.

## RESULTS

The study included 570 participants (114 cases and 456 controls). Most of the participants were male, brown/black, most cases having been diagnosed with spastic and bilateral cerebral palsy (Table 1).

**Table 1.** Characteristics of participants with Cerebral Palsy.

|  | **n** | **%** |
| --- | --- | --- |
| **Sex** |  |  |
| Male | 61 | 53.51 |
| Female | 53 | 46.49 |
| **Race/color** |  |  |
| Yellow | 8 | 7.02 |
| White | 31 | 27.19 |
| Indigenous | 1 | 0.88 |
| Black | 74 | 64.91 |
| **Tipo de Paralisia Cerebral** |  |  |
| Bilateral spastic | 46 | 40.35 |
| Unilateral spastic | 22 | 19.29 |
| Dyskinetic | 2 | 1.75 |
| Ataxic | 3 | 2.63 |
| Uninformed | 41 | 35.96 |
| **Total** | **114** | **100** |

When analyzing the mothers’ socio-demographic characteristics by univariate logistic regression, a significantly higher rate of CP was found in the advanced age group (OR = 3.25 [95% CI 1.88, 5.63]). The stratum of mothers in adulthood was found to be protective (OR = 0.58, [95% CI 0.38, 0.89]). No differences were observed for marital status. Mothers with higher schooling were more likely to have children with CP (OR = 1.66, [95% CI 1.00, 2.75]) (Table 2).

**Table 2.** Absolute and relative frequencies and odds ratio for cerebral palsy according to the sociodemographic characteristics of mothers.

| Sociodemographic characteristics of mothers |  |  |  |  |  |  |  |  |  |
| --- | --- | --- | --- | --- | --- | --- | --- | --- | --- |
|  | Case |  | Control |  | Total |  | OR | IC | p-value |
|  | n | % | n | % | n | % |  |  |  |
| **Advanced age mothers** |  |  |  |  |  |  |  |  |  |
| Yes | 26 | 23% | 38 | 8% | 64 | 11% | 3.25 | (1.88,5.63) | <0.001 |
| No | 88 | 77% | 418 | 92% | 506 | 89% |  |  |  |
| **Lives with his partner** |  |  |  |  |  |  |  |  |  |
| Yes | 47 | 41% | 195 | 43% | 242 | 42% | 0.94 | (0.62,1.42) | 0.767 |
| No | 67 | 59% | 261 | 57% | 328 | 58% |  |  |  |
| **Maternal education** |  |  |  |  |  |  |  |  |  |
| Less than 3 years of study |  |  |  |  |  |  |  |  |  |
| Yes | 9 | 8% | 32 | 7% | 41 | 7% | 1.14 | (0.53,2.45) | 0.746 |
| No | 105 | 92% | 424 | 93% | 529 | 93% |  |  |  |
| **Total** | 114 | 100% | 456 | 100% | 570 | 100% |  |  |  |

Among the prenatal aspects investigated, the presence of congenital anomalies showed a significant effect, with the highest probability of CP occurrence in the univariate analysis (OR = 54.28, [95% CI 12.55, 234.86]). The number of prenatal consultations and the history of other pregnancies did not correlate with differences in CP occurrence (Table 3).

**Table 3.** Absolute and relative frequencies and odds ratio for cerebral palsy according to prenatal characteristics.

|  | Case |  | Control |  | Total |  | OR | IC | p-value |
| --- | --- | --- | --- | --- | --- | --- | --- | --- | --- |
|  | n | % | n | % | n | % |  |  |  |
| **Prenatal consultations** |  |  |  |  |  |  |  |  |  |
| < de 6 consultations | 62 | 54% | 220 | 48% | 282 | 49% | 1.28 | (0.85,1.93) | 0.241 |
| ≥ de 7 consultations | 52 | 46% | 236 | 52% | 288 | 51% |  |  |  |
| **Adequate prenatal care** |  |  |  |  |  |  |  |  |  |
| Yes | 70 | 61% | 264 | 58% | 334 | 59% | 1.15 | (0.76,1.75) | 0.512 |
| No | 44 | 39% | 191 | 42% | 235 | 41% |  |  |  |
| **History of live fetus** |  |  |  |  |  |  |  |  |  |
| Yes | 54 | 47% | 218 | 48% | 272 | 48% | 0.98 | (0.65,1.48) | 0.933 |
| No | 60 | 53% | 238 | 52% | 298 | 52% |  |  |  |
| **History of a dead fetus** |  |  |  |  |  |  |  |  |  |
| Yes | 18 | 16% | 84 | 18% | 102 | 18% | 0.83 | (0.48,1.45) | 0.512 |
| No | 96 | 84% | 372 | 82% | 468 | 82% |  |  |  |
| **Anomalies** |  |  |  |  |  |  |  |  |  |
| Yes | 22 | 19% | 2 | 0.4% | 24 | 4% | 54.28 | (12.55,234.86) | <0.001 |
| No | 92 | 81% | 454 | 99.6% | 546 | 96% |  |  |  |
| **Total** | 114 | 100% | 456 | 100% | 570 | 100% |  |  |  |

\* Adequate prenatal refers to the number of consultations proportional to gestational age.
OR: Odds Ratio; CI: Confidence Interval.

The analysis of the perinatal factors revealed significant differences for low birth weight, gestational age of less than 37 weeks and Apgar lower than 7 at one and five minutes. Multiple gestations, type of delivery and time of birth were not significantly associated with CP occurrence (Table 4).

**Table 4.** Absolute and relative frequencies and odds ratio for cerebral palsy according to perinatal characteristics.

| Characteristics | Case |  | Control |  | Total |  | OR | IC | p-value |
| --- | --- | --- | --- | --- | --- | --- | --- | --- | --- |
|  | n | % | n | % | n | % |  |  |  |
| Type of pregnancy |  |  |  |  |  |  |  |  |  |
| Multiple | 3 | 3% | 16 | 4% | 19 | 3% | 0.74 | (0.21,2.60) | 0.641 |
| Single | 111 | 97% | 440 | 96% | 551 | 97% |  |  |  |
| Type of Delivery |  |  |  |  |  |  |  |  |  |
| Caesarean | 55 | 48% | 212 | 46% | 267 | 47% | 1.07 | (0.71,1.62) | 0.737 |
| Vaginal | 59 | 52% | 244 | 54% | 303 | 53% |  |  |  |
| **Birth time** |  |  |  |  |  |  |  |  |  |
| Night | 53 | 46% | 197 | 43% | 250 | 44% | 1.14 | (0.76,1.72) | 0.527 |
| Day | 61 | 54% | 259 | 57% | 320 | 56% |  |  |  |
| **Low weight** |  |  |  |  |  |  |  |  |  |
| Yes | 38 | 33% | 53 | 12% | 91 | 16% | 3.8 | (2.34,6.16) | <0.001 |
| No | 76 | 67% | 403 | 88% | 479 | 84% |  |  |  |
| **Preterm** |  |  |  |  |  |  |  |  |  |
| Yes | 30 | 26% | 61 | 13% | 91 | 16% | 2.31 | (1.41,3.80) | <0.001 |
| No | 84 | 74% | 395 | 87% | 479 | 84% |  |  |  |
| **Abnormal apgar 1'** |  |  |  |  |  |  |  |  |  |
| Yes | 32 | 28% | 38 | 8% | 70 | 12% | 4.29 | (2.54,7.27) | <0.001 |
| No | 82 | 72% | 418 | 92% | 500 | 88% |  |  |  |
| **Abnormal apgar 5'** |  |  |  |  |  |  |  |  |  |
| Yes | 16 | 14% | 5 | 1% | 21 | 4% | 14.73 | (5.27,41.15) | <0.001 |
| No | 98 | 86% | 451 | 99% | 549 | 96% |  |  |  |
| **Total** | 114 | 110% | 456 | 100% | 570 | 100% |  |  |  |

OR: Odds Ratio; CI: Confidence Interval.

Multivariate logistic regression showed a significant association with birth weight, 5-minute Apgar score, maternal age and congenital anomalies (*P*<0.05) (Table 5).

**Table 5.**
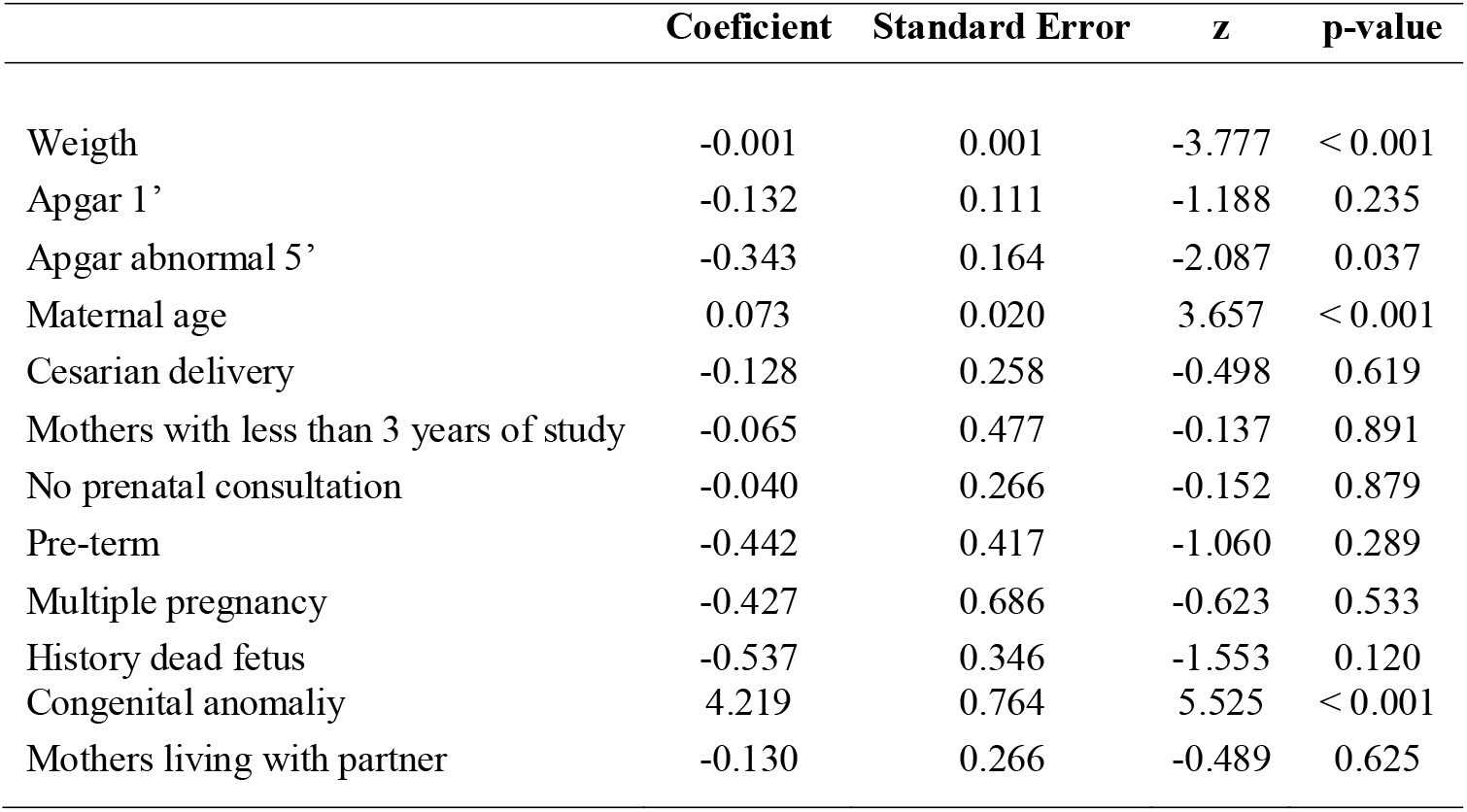
Logistic regression model of risk factors for cerebral palsy.

## COMMENTS

Congenital anomalies had the largest effect in both the univariate and multivariate analyses. Congenital alterations in the development of the CNS of the fetus may determine CP either by themselves, or by increasing the probability of problems occurring during labor, including asphyxia.^19,20^

Microcephaly is the most common congenital defect in cerebral palsy, followed by other changes such as hydrocephalus and anomalies of the corpus callosum.^20^ A study in Norway showed a high proportion of congenital anomalies (25%) among children with late-onset or preterm CP. This type of alteration was associated with a 90-fold increase in risk.^21^

Further studies are needed to analyze the causes of the congenital anomalies that are causing CP in our population: Although some such anomalies have genetic causes, Brazil still faces an epidemic of persistent and preventable transmissible infections, such as syphilis and Zika virus, that can lead to congenital anomalies.^22,23^ Anomalies caused by consanguinity, use of alcohol, tobacco, or other drugs, and unsafe abortion attempts should also be considered.

A low Apgar scores at one and five minutes stood out as an important factor predicting CP, with alterations at five minutes leading to a 14.73 times greater chance of CP. This result was confirmed by the significance of this factor also in multivariate analysis. Other studies found similar results,^19,24–26^ one of them reporting a 13.2-fold higher risk of CP for low Apgar scores.^19^ The authors report that an Apgar score less than 7 at one or five minutes increased the probability of occurrence of CP, especially in preterm infants.

Low Apgar scores should not be considered the cause of CP, because the score is a composite of clinical signs, and a reflection of neonatal cardiorespiratory function that may reflect an acute or chronic fetal impairment. However, these scores are used as indirect manifestations of asphyxia at birth.^19,25,27^ Although low Apgar scores can have other possible causes, researchers believe that they could be considered a marker of causal factors underlying cerebral palsy and related deficiencies.^27,28^

Some evidence has been produced indicating that asphyxia at birth may not be the main cause of CP, and that prenatal factors have a larger role in both preterm and full-term children.^20^ However, neonatal asphyxia is an event of great prevalence in Brazil. A study revealed that 41% of fetal deaths are caused by intrapartum asphyxia, even in hospital deliveries.^22^

Our results indicate that intrapartum events, such as asphyxiation, can play a significant role, second only to that of congenital anomalies, in increasing the probability of CP.

There is a 3.8 times greater chance of CP in infants below 2500 g, and the association remains significant even after multivariate analysis adjustments. These findings are reiterated in the international literature, which considers low birth weight as one of the main risk factors. Several authors found a weight below 2500 g to be a perinatal risk factor.^19,27,29,0^

Preterm birth also increased the probability of CP in our population. Similar data were found in other studies, that found a strong association with this risk factor.^16,19,24– 26,31,32^ However, it is important to note that CP can affect a large proportion of full-term babies, since a Swiss study revealed that 60% of all children with CP were born at the appropriate gestational age.^33,34^ In a survey conducted in China, gestational age between 37 and 41 weeks at birth was found to be protective against CP.^16^

Improvements in health care in the last decades led to increased survival of preterm infants, which however may coexist with permanent neurological sequelae. We emphasize the importance of better investigating the causes of preterm births in the Brazilian context, to improve prenatal and neonatal care to avoid neurological injuries and deficiencies.

Since the year 2000, the prevalence of low birth weight in Brazil has remained stable at the level of 8%, notwithstanding the growth in the number of preterm births. According to a survey, the majority of preterm newborns in Brazil have gestational age between 34 and 36 weeks, and weigh more than 2500 grams.^22^

We propose the hypothesis that in the study population, low-weight and preterm infants are more exposed to events that cause CP-inducing damage, including anoxia. It would be important to conduct further studies to verify the association of these causes with CP in Brazil.

Among the socio-demographic aspects, advanced maternal age was associated with CP. Another study that found similar results argues that older women are highly vulnerable to obesity, gestational diabetes, gestational hypertension, and other conditions that increase the risk of preterm birth, low birth weight, fetal distress, SGA, and other abnormal perinatal outcomes.^16^ The cohabitation of the mother and her companion did not show any association with CP in our population, but another study in Sweden found it to be a risk factor.^33^

One of the prerogatives of the Brazilian health system is universal access to prenatal health monitoring in primary care, and to diagnostic and therapeutic support services. The lack of significant association between prenatal follow-up and CP must be carefully interpreted, and possibly further investigated in future studies. We can formulate two hypotheses in this regard: (1) Prenatal quality analysis would be a better indicator, since the coverage is practically universal, and the number of consultations does not necessarily reflect the health status of women. (2) Unwanted outcomes are due to childbirth itself.

A literature review shows that despite high prenatal coverage in Brazil, this type of health care is poorly integrated into hospital care. The author cites data from a survey conducted in the metropolitan region of Rio de Janeiro, showing that one out of three women in labor had to seek more than one maternity hospital to be hospitalized.^22^

Vaginal or cesarean delivery, and delivery during the day or night shift, did not show a significant association with CP. Data from another case-control study show similar results.^19^ The increase in the rate of cesareans in the last decades did not decrease CP rates.^20^ Therefore, the increase in surgical interventions cannot be justified by the need to decrease the risk of CP.

This study had some limitations worth highlighting, even though they did not compromise the research design. Some variables related to the prenatal and perinatal periods, that are known risk factors according to the literature, were not used since they are not present in the SINASC records. For instance, it was not possible to access data on maternal infectious or chronic diseases, use of prenatal and perinatal drugs, consanguinity, or harmful habits such as smoking, alcoholism and the use of other drugs. Many events that result in CP may also relate to these variables: Therefore we encourage further research to be conducted in Brazil following this perspective, including differentiating how risk factors determine different types of CP.

The main advantage of this study consisted in its population-based design, which guarantees greater consistency of the results. The cases were gathered from all over the city, avoiding possible selection biases. Increasing our knowledge about gestation, delivery, and birth of children with CP can improve our response capacity, and reduce the causes of CP and the damage it inflicts.

In conclusion, the main prenatal and perinatal factors associated with cerebral palsy in our population were congenital anomalies, low Apgar scores, low birth weight, reduced gestational age, and advanced mother age. The perinatal period was associated to more risk factors, demanding a deeper study of causes and preventive measures. The results of this study add to the existing knowledge on the risk factors for cerebral palsy in Brazil, and can serve as the basis for comparisons with data from other countries.

## Data Availability

I declare that the responsible researcher and the research team undertake to make the data available for eventual needs without identifying the participants.

## Notes

Conflict of Interest Statement: The authors have no problems of interest to this article namely disclosed

### Competing Interest Statement

The authors have declared no competing interest.

### Funding Statement

The authors have no relevant financial relationships for this article to disclose.

### Author Declarations

The research was approved by the Federal University of Sergipe Research Ethics Committee (opinion number 1.177.455) which abides by the ethical principles of the Brazilian National Council of Ethics in Research (Conselho Nacional de Etica em Pesquisa do Brasil - CONEP) and the Declaration of Helsinki.

## REFERENCES

1. Rosenbaum P, Paneth N, Leviton A, Goldstein M, Bax M, Damiano D, et al. A report: The definition and classification of cerebral palsy April 2006. Dev Med Child Neurol. 2007;49(SUPPL.109):8–14. doiorg/10.1111/j.1469-8749.2007.tb12610.x. PMID: 17370477.

2. Graham HK, Rosenbaum P, Paneth N, Dan B, Lin JP, Damiano DL, Becher JG, Gaebler-Spira D, Colver A, Reddihough DS, Crompton KE, Lieber RL. Cerebral palsy. Nat Rev Dis Primers. 2016 Jan 7;2:15082. doi: 10.1038/nrdp.2015.82. doi:10.1038/nrdp.2015.82. PMID: 27188686.

3. Pakula AT, Van Naarden Braun K, Yeargin-Allsopp M. Cerebral Palsy: Classification and Epidemiology. Phys Med Rehabil Clin N Am. 2009;20(3):425–52. dx.doi.org/10.1016/j.pmr.2009.06.001. PMID: 19643346.

4. Mcintyre S, Taitz D, Keogh J, Goldsmith S, Badawi N, Blair E. A systematic review of risk factors for cerebral palsy in children born at term in developed countries. Dev Med Child Neurol. 2013 Jun;55(6):499–508. doi.wiley.com/10.1111/dmcn.12017. PMID: 23181910.

5. Colver A, Fairhurst C, Pharoah POD. Cerebral palsy. Lancet. 2014 Apr;383(9924):1240–9. doi: 10.1016/S0140-6736(13)61835-8. PMID: 24268104.

6. Saadi HR, Sutan R, Dhaher AM, Alshaham SA. Maternal and foetal risk factors of cerebral palsy among Iraqi children. A case control study. Open J Prev Med. 2012;02(03):350–8. DOI:10.4236/ojpm.2012.23051

7. Oskoui M, Coutinho F, Dykeman J, Jetté N, Pringsheim T. An update on the prevalence of cerebral palsy: A systematic review and meta-analysis. Dev Med Child Neurol. 2013 Jun;55(6):509–19. doi: 10.1111/dmcn.12080. PMID: 23346889

8. Walstab J, Bell R, Reddihough D, Brennecke S, Bessell C, Beischer N. Antenatal and intrapartum antecedents of cerebral palsy: a case-control study. Aust N Z J Obstet Gynaecol. 2002;42(2):138–46. doi: 10.1111/j.0004-8666.2002.00138.x. PMID: 12069139.

9. Himmelmann K, Ahlin K, Jacobsson B, Cans C, Thorsen P. Risk factors for cerebral palsy in children born at term. Acta Obstet Gynecol Scand. 2011;90(10):1070–81. doi: 10.1111/j.1600-0412.2011.01217.x. PMID: 21682697.

10. Sellier E, Platt Mjmj, Andersen Glgl, Krägeloh-Mann I, De La Cruz J, Cans C, et al. Decreasing prevalence in cerebral palsy: A multi-site European population-based study, 1980 to 2003. Dev Med Child Neurol. 2016;58(1):85–92. doi: 10.1111/dmcn.12865. PMID: 26330098.

11. Petridou E, Koussouri M, Toupadaki N, Papavassiliou A, Youroukos S, Katsarou E, et al. Risk factors for cerebral palsy: A case-control study in Greece. Scand J Public Health. 1996;24(1):14–26. doi: 10.1177/140349489602400104. PMID: 8740872.

12. Serdaro[lu A, Cansu A, Özkan S, Tezcan S. Prevalence of cerebral palsy in Turkish children between the ages of 2 and 16 years.Dev Med Child Neurol. 2006;48(6):413–6. doi: 10.1017/S0012162206000910. PMID: 16700929.

13. Oztürk A, Demirci F, Yavuz T, Yildiz S, Değirmenci Y, Döşoğlu M, et al. Antenatal and delivery risk factors and prevalence of cerebral palsy in Duzce (Turkey). Brain Dev. 2007 Jan;29(1):39–42. doi: 10.1016/j.braindev.2006.05.011. PMID: 16824718.

14. Daher S, El-khairy L.Risk factors for cerebral palsy in Palestinian children[: a case-control study. Lancet. 2013;382:S8. dx.doi.org/10.1016/S0140-6736(13)62580-5

15. Donald KA, Neuro MP, Samia P, Neuro MP, Kakooza-mwesige A, Bearden D. Pediatric Cerebral Palsy in Africa[: A Systematic Review. Semin Pediatr Neurol. 2014;21(1):30–5. http://dx.doi.org/10.1016/j.spen.2014.01.001

16. Gao J, Zhao B, He L, Sun M, Yu X, Wang L. Risk of cerebral palsy in Chinese children: A N:M matched case control study. J Paediatr Child Health. 2017;53(5):464–9. doi: 10.1111/jpc.13479. Epub 2017 Jan 30. PMID: 28134474.

17. Gladstone M. A review of the incidence and prevalence, types and aetiology of childhood cerebral palsy in resource-poor settings. Ann Trop Paediatr. 2010;30(3):181 96. doi/full/10.1179/146532810×12786388978481

18. World Health Organization. International Statistical Classification of Diseases and Related Health Problems [Internet]. 10th ed. Genebra: World Health Organization; 2016. Available from: http://apps.who.int/classifications/icd10/browse/2016/en

19. Gurbuz A, Karateke A, Yilmaz U, Kabaca C. The role of perinatal and intrapartum risk factors in the etiology of cerebral palsy in term deliveries in a Turkish population. J Matern Neonatal Med. 2006;19(3):147–55. doi: 10.1080/14767050500476212. PMID: 16690507.

20. Nelson KB, Blair E. Prenatal Factors in Singletons with Cerebral Palsy Born at or near Term. N Engl J Med. 2015 Sep 3;373(10):946–53. doi: 10.1056/NEJMra1505261. PMID: 26332549.

21. Jystad KP, Strand KM, Bjellmo S, Lydersen S, Klungsöyr K, Stoknes M, et al. Congenital anomalies and the severity of impairments for cerebral palsy. Dev Med Child Neurol. 2017;59(11):1174–80. doi.wiley.com/10.1111/dmcn.13552. PMID: 28967231.

22. Victora CG, Aquino EM, Do Carmo Leal M, Monteiro CA, Barros FC, Szwarcwald CL. Maternal and child health in Brazil: Progress and challenges. Lancet. 2011;377(9780):1863–76. dx.doi.org/10.1016/S0140-6736(11)60138-4

23. Marcondes CB, Ximenes M de FF de M. Zika virus in Brazil and the danger of infestation by Aedes (Stegomyia) mosquitoes. Rev Soc Bras Med Trop. 2016;49(1):4-10. doi.org/10.1590/0037-8682-0220-2015

24. Jacobsson B, Hagberg G, Hagberg B, Ladfors L, Niklasson A, Hagberg H. Cerebral palsy in preterm infants: a population-based case-control study of antenatal and intrapartal risk factors. Acta Paediatr. 2002;91(8):946–51. doi.wiley.com/10.1080/080352502760148685

25. O[Callaghan ME, MacLennan AH, Gibson CS, McMichael GL, Haan E a., Broadbent JL, et al. Epidemiologic Associations With Cerebral Palsy. Obstet Gynecol. 2011 Sep;118(3):576–82. Sep;118(3):576-82. doi: 10.1097/AOG.0b013e31822ad2dc. PMID: 21860286

26. Stelmach T, Pisarev H, Talvik T. Ante- and Perinatal Factors for Cerebral Palsy: Case-Control Study in Estonia. J Child Neurol. 2005 Aug 2;20(8):654–61. doi/10.1177/08830738050200080401. PMID: 16225810

27. Lie KK, Grøholt EK, Eskild A. Association of cerebral palsy with Apgar score in low and normal birthweight infants: Population based cohort study. BMJ. 2010;341(7777):817. doi: 10.1136/bmj.c4990. PMID: 20929920.

28. Persson M, Razaz N, Tedroff K, Joseph KS, Cnattingius S. Five and 10 minute Apgar scores and risks of cerebral palsy and epilepsy: population based cohort study in Sweden. BMJ. 2018 Feb 7;360:k207. doi/10.1136/bmj.k207. PMID: 29437691

29. Yeargin-Allsopp M, Van Braun KN, Doernberg NS, Benedict RE, Kirby RS, Durkin MS. Prevalence of cerebral palsy in 8-year-old children in three areas of the united states in 2002: A multisite collaboration. Pediatrics. 2008;121(3):547–54. doi: 10.1542/peds.2007-1270. PMID: 18310204

30. Hirvonen M, Ojala R, Korhonen P, Haataja P, Eriksson K, Gissler M, et al. Cerebral Palsy Among Children Born Moderately and Late Preterm. Pediatrics. 2014;134(6):e1584–93. doi: 10.1542/peds.2014-0945. PMID: 25422011

31. Moster D, Wilcox AJ, Vollset SE, Markestad T, Lie RT. Cerebral palsy among term and postterm births. JAMA - J Am Med Assoc. 2010;304(9):976–82. doi: 10.1001/jama.2010.1271. PMID: 20810375

32. Ahlin K, Himmelmann K, Hagberg G, Kacerovsky M, Cobo T, Wennerholm UB, et al. Non-infectious risk factors for different types of cerebral palsy in term-born babies: A population-based, case-control study. BJOG An Int J Obstet Gynaecol. 2013;120(6):724–31. doi: 10.1111/1471-0528.12164. PMID: 23418811.

33. Himmelmann K, Uvebrant P. The panorama of cerebral palsy in Sweden part XII shows that patterns changed in the birth years 2007–2010. Acta Paediatr Int J Paediatr. 2018;107(3):462–8. doi: 10.1111/apa.14147. PMID: 29121418

